# Reduced expression of COVID-19 host receptor, *ACE2* is associated with small bowel inflammation, more severe disease, and response to anti-TNF therapy in Crohn’s disease

**DOI:** 10.1101/2020.04.19.20070995

**Authors:** Alka A. Potdar, Shishir Dube, Takeo Naito, Gregory Botwin, Talin Haritunians, Dalin Li, Shaohong Yang, Janine Bilsborough, Lee A. Denson, Mark Daly, Stephan R. Targan, Phillip Fleshner, Jonathan Braun, Subra Kugathasan, Thaddeus S. Stappenbeck, Dermot P.B. McGovern

## Abstract

Angiotensin-Converting Enzyme 2 (*ACE2*) has been identified as the host receptor for SARS-coronavirus 2 (SARS-CoV-2) which has infected millions world-wide and likely caused hundreds of thousands of deaths. Utilizing transcriptomic data from four cohorts taken from Crohn’s disease (CD) and non-inflammatory bowel disease (IBD) subjects, we observed evidence of increased *ACE2* mRNA in ileum with demographic features that have been associated with poor outcomes in COVID-19 including age and raised BMI. *ACE2* was downregulated in CD compared to controls in independent cohorts. Within CD, *ACE2* expression was reduced in inflamed ileal tissue and also remarkably, from un-involved tissue in patients with a worse prognosis in both adult and pediatric cohorts. In active CD, small bowel *ACE2* expression was restored by anti-TNF therapy particularly in anti-TNF responders. Collectively our data suggest that *ACE2* downregulation is associated with inflammation and worse outcomes in CD.

## Introduction

As of April 18^th^ 2020 approximately two and a quarter million people worldwide have confirmed Coronavirus disease 2019 (COVID-19) infection with current (and likely conservative) estimates implicating the virus in over 150,000 deaths. COVID-19, caused by infection with SARS-coronavirus 2 (SARS-COV-2), most commonly presents with respiratory symptoms. However, recent reports have suggested that patients can frequently have both respiratory and GI symptoms (predominantly diarrhea and nausea) and in a proportion of patients GI symptoms may be the sole complaint [1-3]. There has also been concern that detection of the virus in stool may implicate the fecal-oral route as an important mode of transmission.

There is very significant variation in outcomes from COVID-19 with the majority having mild symptoms, a minority with respiratory complications, and a small percentage dying as a consequence of secondary cytokine storm or superimposed infection. Increasing age, male gender, smoking, co-morbidities, and an elevated body mass index (BMI) have all been implicated in increased morbidity and mortality but it is likely that other factors also contribute to the variability in response [4-7]. There is understandable interest and concern in the role that immunosuppressive medications commonly used in immune-mediated diseases may have on the susceptibility and natural history of COVID-19.

Angiotensin-Converting Enzyme 2 (*ACE2*) is a putative receptor for SARS-COV-2 entry into human cells. Other molecules that interact with *ACE2* and are plausible candidates in COVID-19 biology include: the transmembrane serine protease (*TMPRSS2*) that helps prime SARS-COV-2 spike protein for host cell entry [8]); the *ACE2* paralog in the renin–angiotensin– aldosterone system (RAAS), angiotensin I converting enzyme (*ACE*); and the solute carrier family 6 member 19 (*SLC6A19*), expression of which is dependent on *ACE2* [9]. The expression of *ACE2* is altered in fibrotic pulmonary disease and in the lung tissue of smokers [5, 10, 11]. *ACE2* is abundantly expressed in small bowel (SB) compared to other tissues including whole blood [12]. Our aim was to determine factors including inflammation and drug treatment, that influenced *ACE2* expression in the SB of Crohn’s disease (CD) patients and non-IBD (inflammatory bowel disease) controls and to investigate shared disease biology between IBD and COVID-19.

## Methods

### Tissue Samples and Study Subjects

We investigated association of *ACE2* mRNA with age at collection, gender, smoking, BMI, diagnosis, CD sub-phenotypes and cytokine levels in 4 independent transcriptomic datasets of SB gene expression contingent on availability of meta-data for each cohort (see Table 1). Three of these cohorts have been described previously. In all 4 cohorts the small bowel specimens were taken from macroscopically normal appearing tissue.

**Table 1.** Details of the four transcriptomic cohorts used to study association with available demographics and disease status. ^*^See text for details.

| Name | Platform (expression) | GEO /ArrayExpress | Numbers |  |  | Meta-data availability (N) |  |  |  |  |  |
| --- | --- | --- | --- | --- | --- | --- | --- | --- | --- | --- | --- |
|  |  |  | non-IBD | CD | UC | Age at collection (N) | Gender | BMI at surgery | Smoking | Disease status | CD sub-phenotypes |
| SB139 | Microarray (log2) | GSE120782 |  | 139 |  | Yes (139) | Yes(139) | No | Yes(127) | Yes(139) | Yes* |
| WashU | RNA-seq (FPKM) | E-MTAB-5783 | 32 | 38 |  | Yes (70) | Yes(55) | Yes(66) | Yes(34) | Yes(70) | No |
| Cedars 100 | RNA-seq (RPKM) | Unpublished |  | 100 |  | Yes (99) | Yes(100) | No | Yes (78) | Yes(100) | Yes* |
| RISK | RNA-seq (RPKM) | GSE57945 | 42 | 218 | 62 | Yes (322) | Yes(322) | No | N/A | Yes(322) | Yes* |

The ‘SB139’ [13] dataset was generated using whole Human Genome 4×44k Microarrays (Agilent) from formalin fixed paraffin embedded (FFPE) tissue taken from the unaffected margin of SB tissue resected during ileo-cecal or SB resection for complicated CD. Median age at time of surgery, which were all performed at Cedars-Sinai Medical Center, Los Angeles, was 32 years. The ‘WashU’ dataset [14] was generated by RNA-seq and similarly was generated from FFPE tissue from the unaffected proximal margin of resected CD tissues and also from FFPE from non-IBD control subjects. These subjects had a median age of 51 years at time of surgery which were all performed at the University of Washington, St Louis. The ‘RISK’ [15, 16] dataset was generated by RNA-seq from ileal biopsies taken from pediatric subjects in a CD inception cohort from multiple centers across North America (median age at time of biopsy 12 years). Being an inception cohort the age of diagnosis and age at specimen collection are the same. The CD subjects in RISK cohort were divided into two groups: those that had no small bowel/ileal disease (cCD) and those where the ileum was involved (iCD). The ‘Cedars100’ [17] dataset has not been previously published but similarly, utilized FFPE from un-involved proximal resection margins from complicated CD surgeries (performed at Cedars-Sinai Medical Center) and transcriptomics were generated by RNA-seq. All study subjects in SB139 and Cedars100 were CD; the WashU cohort consisted of CD and non-IBD controls and RISK cohort is a mix of CD, UC, and non-IBD controls.

In addition we looked at the effect of drug exposure on SB *ACE2* expression from two clinical trials of biologic therapies commonly used in CD: infliximab (IFX cohort, GSE16879) [18] and ustekinumab (UST cohort, GSE100833) [19]). Briefly, the transcriptomics for the IFX cohort were generated using Affymetrix Human Genome U133 Plus 2.0 microarray platform using biopsies from inflamed mucosa (n=61 IBD subjects) before and 4-6 weeks after first infliximab infusion and in normal mucosa from 12 control patients (6 colon and 6 ileum). The patients were classified as responders/non-responders for treatment based on endoscopic and histologic findings at 4-6 weeks after infliximab induction treatment. We only focused on SB ileal transcriptomics from IFX cohort for the purpose of this study.

The UST cohort consists of microarray (Affymetrix HT HG-U133+ PM Array Plate) transcriptomics of human blood and intestinal biopsy samples from a phase 2b, double-blind, placebo-controlled study of ustekinumab in CD [19]. The cohort contained gene expression on 329 biopsies from multiple regions in the intestine of 87 Crohn’s disease subjects. For consistency, we only focused on SB ileal transcriptomics for the purpose of this study. Response outcomes to ustekinumab were not available.

### Study approval

For SB139 and Cedars100 cohorts, tissue samples and genetic data were obtained by the Material and Information Resources for Inflammatory and Digestive Diseases [MIRIAD] IBD Biobank after the patients’ informed consent and approval by the IRB of the Cedars-Sinai Medical Center [protocol #3358]. The other datasets were all previously published, and details of approvals can be found in the publications [14-16, 18, 19].

### Transcriptomics data generation and processing

The Genome Technology Access Center at Washington University (St Louis, MO) generated datasets in the SB139, WashU and Cedars100 cohorts. The methods used to generate microarray SB139 cohort data have been previously described here [13]. For the WashU cohort, RNA-seq library preparation, sequencing and read alignment was performed and sequencing done on an Illumina HiSeq2000 SR42 (Illumina, San Diego, CA) using single reads extending 42 bases.

For the Cedars100 cohort, total RNAs were processed with Sigma Seqplex to create amplified ds-cDNA, followed by traditional Illumina library preparation with unique dual indexing. 100 libraries were run on NovaSeq6000, S2 flow cell, using single-end 100 base reads. The run generated approximately 4.2B reads passing filter, thus an average of 42 million reads per library were generated. The data generation methods for other three cohorts (RISK, IFX, UST) have been published previously[15, 16, 18, 19].

Table 1 shows the GEO accession numbers of the two of the three published cohorts used in the study. The dataset for the WashU cohort can be accessed on ArrayExpress. The methods used to process microarray data from SB139 cohort have been previously described [13]. The pipeline used for RNA-seq data processing and normalizing for the Cedars100 cohort was similar to the one used for the WashU cohort as previously described[14]. For Cedars100, RNA-seq data was normalized and resultant RPKM values were generated for analysis while for WashU normalized data were generated in FPKM. The methods used to process the RNA-seq data from RISK cohort have also described previously [15, 16]. The data processing methods for IFX cohort and UST cohort are described in prior publications [18, 19].

Normalized processed data for some cohorts (RISK, IFX and UST) were downloaded using accession numbers available at GEO in series matrix files which were cleaned and annotated with geneids. Clean, processed data for SB139, Cedars100 and WashU along with respective meta-data was available in-house.

### Clinical and Demographic Data

Meta-data available for the different transcriptomics cohorts used is compiled in Table 1. Clinical phenotype data available for SB139 included age at collection, gender, smoking status, disease status, CD sub-phenotypes and disease recurrence after surgery. The Cedars100 cohort included age at collection, gender, smoking status, disease status and CD sub-phenotypes.

For the ‘WashU cohort, data were extracted from the clinical charts and includes age at collection, gender, disease status, smoking and BMI at surgery. The ‘RISK’ cohort is a pediatric inception cohort and data were collected prospectively. The transcriptomics dataset and some meta-data for RISK cohort were downloaded from NCBI (GEO/SRA) using GSE57945 accession number (age at collection, gender and disease diagnosis (including information for involved versus unaffected CD)) and complication data were available from the prospective follow up.

The ‘CD sub-phenotypes’ meta-data in Table 1 includes severe versus mild refractory in SB139, involved versus un-involved in RISK and disease complication (B1=inflammatory; B2=stricturing, B3=penetrating in SB139, Cedars100 and RISK). Meta-data for IFX (GSE16879) and UST (GSE100833) cohorts was downloaded from their respective GEO accession numbers.

### Methods for datasets downloaded via GEO

Platform annotation, normalized gene expression, and phenotype meta-data were extracted using the R package GEOquery (GEO2R library). The phenotype meta-data table was used to identify categories such as tissue type (non-involved/inflamed terminal ileum biopsy tissue samples), disease status (Control, CD, UC), time points (defined as week 0 and week 6) for treatment, treatment type, etc. as available per cohort.

### Univariate and Multivariate model fits

Univariate models were fitted with *ACE2* as response and each available demographic data (age, gender, BMI at surgery, smoking status) as a predictor in each cohort. A similar pipeline was followed for clinical predictors such as disease status, CD severity sub-groups, recurrence, and treatment where available in a given cohort. This was followed by fitting multivariate models with *ACE2* expression as response and all available predictors within each cohort.

In the WashU and RISK cohorts, multivariate models were also fitted for expression of other COVID-19 relevant genes such as *ACE, TMPRSSS2* and *SLC6A19*. We examined the relationship between *ACE2* expression and disease recurrence (only available in SB139) through a multivariate model with age, gender and first two principal components in genotype data calculated using genetic data published previously [13]. We also examined association of *ACE2* with CD disease behavior B1, B2 and B3 (available in SB139, Cedars100 and RISK) using age and gender as covariates.

### Statistical tools

Statistical package glm in R (version 3.5.1) was used to perform univariate and multivariate associations with a p < 0.05 cutoff for statistical significance. In some cases, GraphPad Prism 7 (GraphPad, La Jolla, CA) was also used to perform statistical analysis with appropriate parametric or nonparametric statistical tests applied.

### ACE2 gene co-expression analysis

Co-expression analysis of *ACE2* with 54 genes of interest involved in either IBD pathogenesis [20] or high probability SARS CoV-2 virus-host protein-protein interaction [21] was performed using the SB139 and Cedars100 cohorts. Genomic annotations for candidate genes of interest were extracted at the probe/transcript level from the platform annotation file for SB139 [13] and Cedars100 [R based GenomicFeatures package in Bioconductor]. The statistical package glm was used to fit a multivariate linear regression model on the gene pairs and included covariates, such as age at collection and gender (when available) with a p < 0.05 cutoff for statistical significance. The full list of genes examined in the co-expression analysis are available in Table S1.

## Results

### Differences in *ACE2* gene expression with age, BMI, disease, smoking and gender

#### Univariate associations

We examined *ACE2* mRNA expression by age of the subject at the time of specimen collection when available. The expression of the most abundantly expressed *ACE2* transcript isoform (ENST00000252519) was associated with age at collection in the WashU cohort (Figure 1a) with higher expression being associated with older age at collection, as observed in CD and non-IBD controls. The association with age trended towards significance in the pediatric RISK cohort (Figure 1b). We did not see statistically significant association with age in the microarray based SB139 cohort (Figure S1) as well as Cedars100 cohort. Combining data from RNA-seq platform-based WashU, Cedars100 and RISK cohorts to generate fold-change of *ACE2* gene expression with respect to the house-keeping gene *GAPDH* in the respective cohorts, validated the positive correlation of age at specimen collection with *ACE2* (Figure 1c). In the WashU cohort, we observed a strong association of *ACE2* expression with BMI in both CD and non-IBD controls, with subjects with higher BMI having elevated *ACE2* expression (p<0.0001, linear regression) (Figure 2).

**Figure 1:**
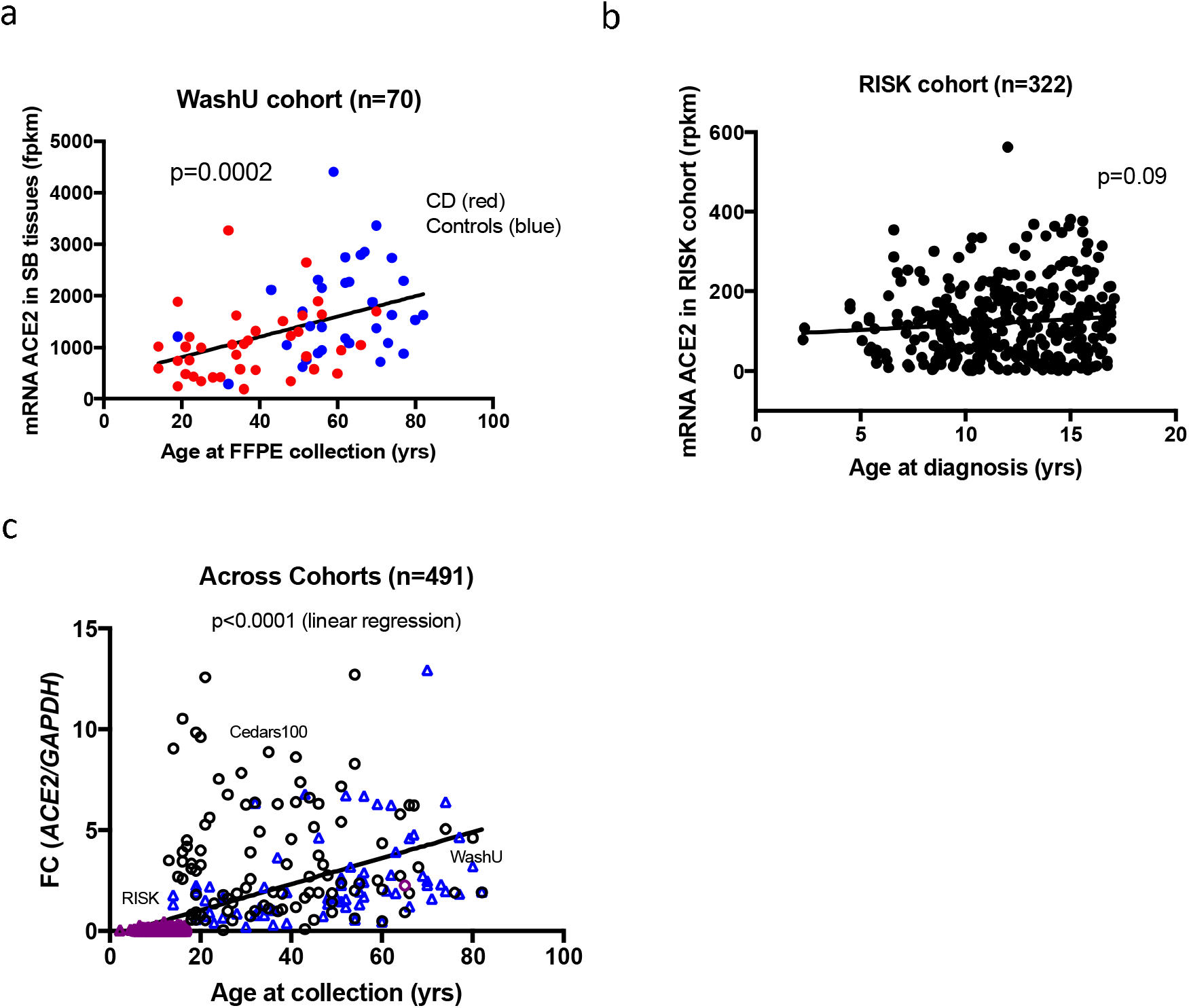
Association of *ACE2* with age at collection; a) WashU cohort; b) RISK cohort c) Across combination of RNA-seq platform-based three SB cohorts (RISK, Cedars100 and WashU).

**Figure 2:**
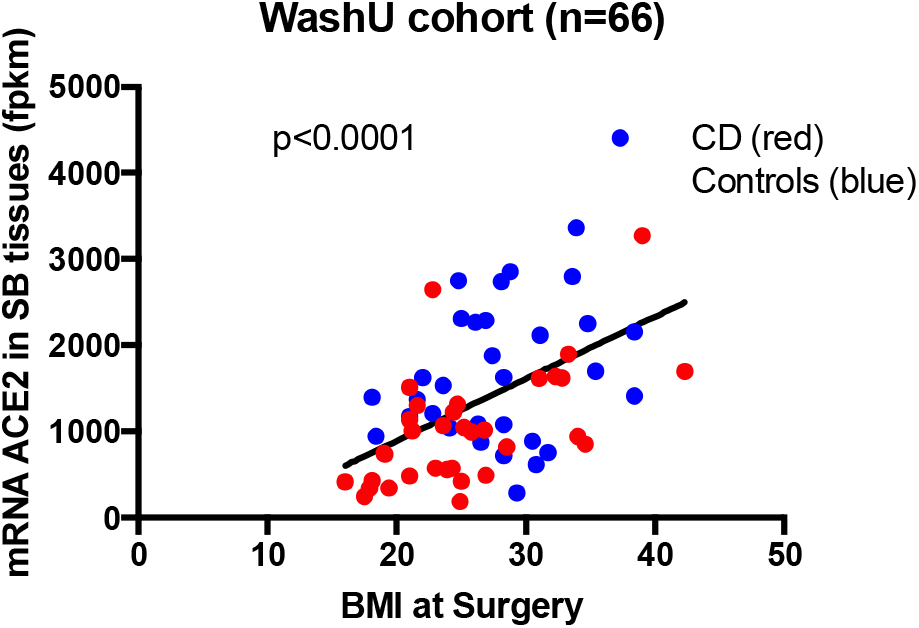
Association of *ACE2* with BMI in WashU cohort using linear regression.

We did not see significant association with gender in SB139, WashU and RISK cohorts (Figure S1, Table 2, 3 and 4). However, we did observe higher ileal expression of *ACE2* in females in the Cedars100 cohort (Figure 3a), consistent with similar observations in GTEx [12]. We did not find statistical association of smoking with *ACE2* expression in any of the adult cohorts (Table 2 and Figure S1) although there was a suggestive trend towards higher expression, in the Cedars100 cohort (Figure 3b) (p=0.08).

**Table 2:** Univariate and multivariate models of associations of *ACE2* mRNA expression in WashU cohort. Comparison of disease status performed with CD as reference and for gender with male as reference.

| Response : *ACE2* (FPKM) | Univariate |  |  | Multivariate |  |  | Multivariate |  |  | Multivariate |  |  |
| --- | --- | --- | --- | --- | --- | --- | --- | --- | --- | --- | --- | --- |
|  | Beta | P | N | Beta | P | N | Beta | P | N | Beta | P | N |
| BMI at surgery | 71.99 | 0.000017 | 66 | 51.37 | 0.002 | 51 |  |  |  |  |  |  |
| Age at collection | 19.71 | 0.000176 | 70 | 5.65 | 0.420 | 51 | 9.42 | 0.167 | 55 | 13.49 | 0.036 | 70 |
| Disease status (Control) | 684.30 | 0.000515 | 70 | 487.7 | 0.052 | 51 | 550.6 | 0.039 | 55 | 369.78 | 0.120 | 70 |
| Gender (Female) | -5.56 | 0.979007 | 55 | 78.47 | 0.672 | 51 | -30.08 | 0.873 | 55 |  |  |  |
| Smoking (Yes) | 146.90 | 0.523000 | 34 |  |  |  |  |  |  |  |  |  |

**Table 3:**
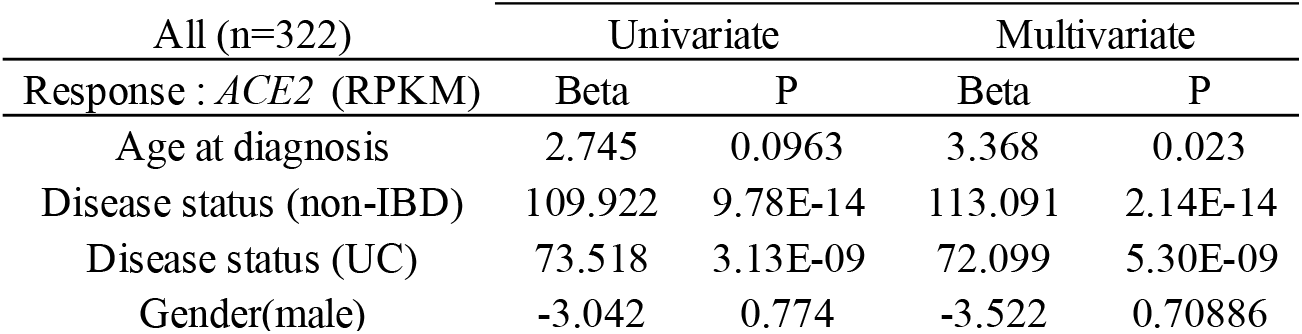
Univariate and multivariate models of associations of *ACE2* mRNA expression in the RISK cohort (numbers: total = 322; non-IBD controls =42, UC = 62, CD = 218). Comparison of disease status done with CD as reference and for gender with female as reference.

**Table 4:**
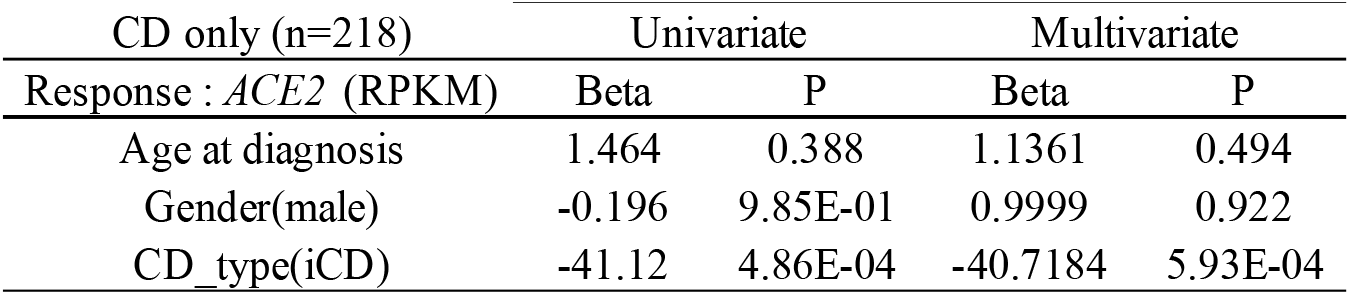
Univariate and multivariate models of associations of *ACE2* mRNA expression within CD subjects in RISK cohort (numbers: total = 218; iCD = 163, cCD = 55). Comparison of disease status performed with cCD as reference and for gender with female as reference.

**Figure 3:**
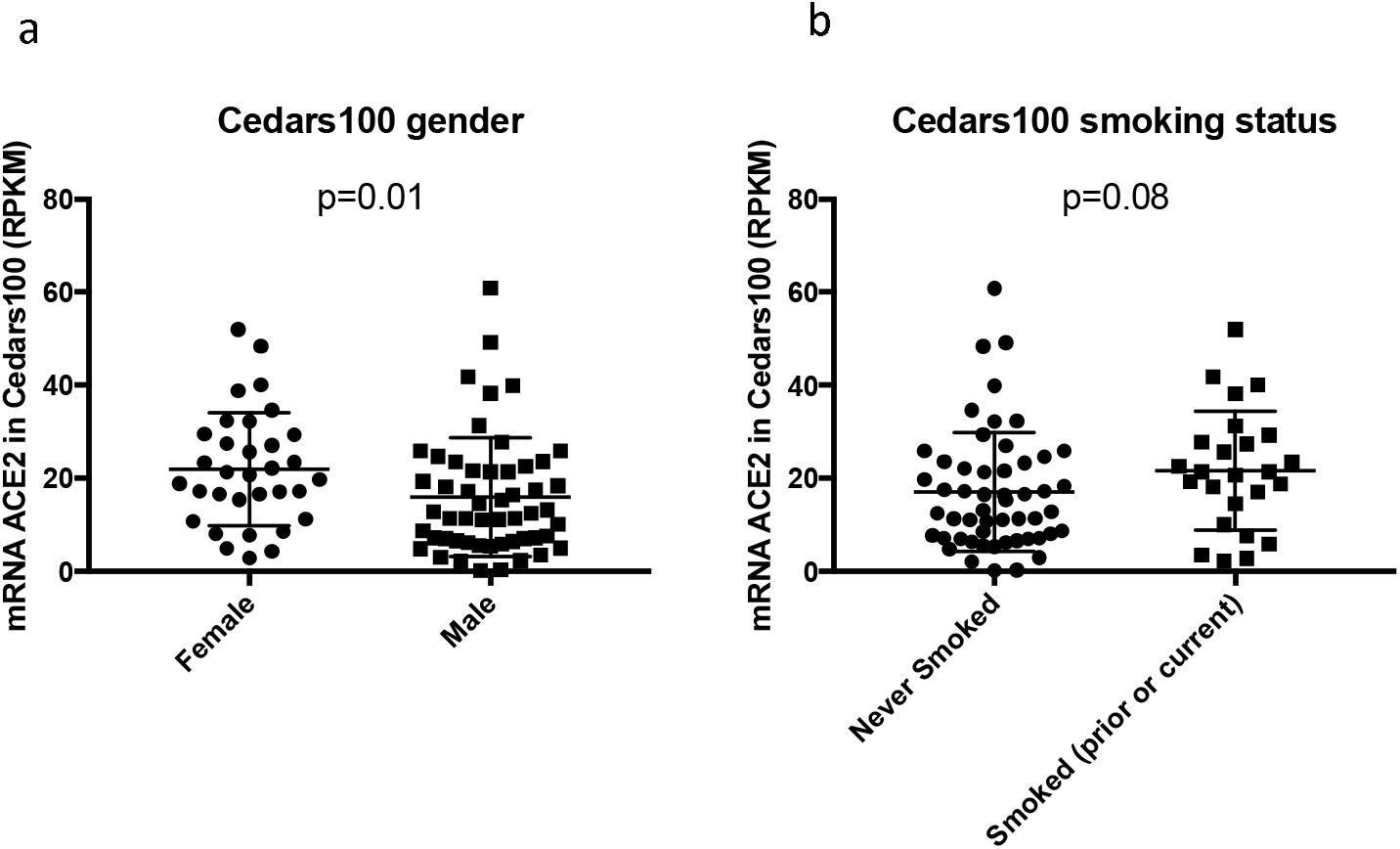
*ACE2* levels and demographics a) Univariate association of *ACE2* in Cedars100 cohort with gender indicating lower expression in males (p=0.01, Mann-Whitney test); b) with smoking status indicating higher expression if prior or current smoker (p=0.08, Mann-Whitney test).

Data from non-IBD controls for comparison were only available for the WashU and RISK cohorts. In the WashU cohort (Figure 4a), *ACE2* expression was lower in CD compared to non-IBD controls (p=0.0004). A univariate model with disease status as the predictor, was statistically significant for lower *ACE2* expression in CD versus control in the WashU cohort (Table 2).

**Figure 4:**
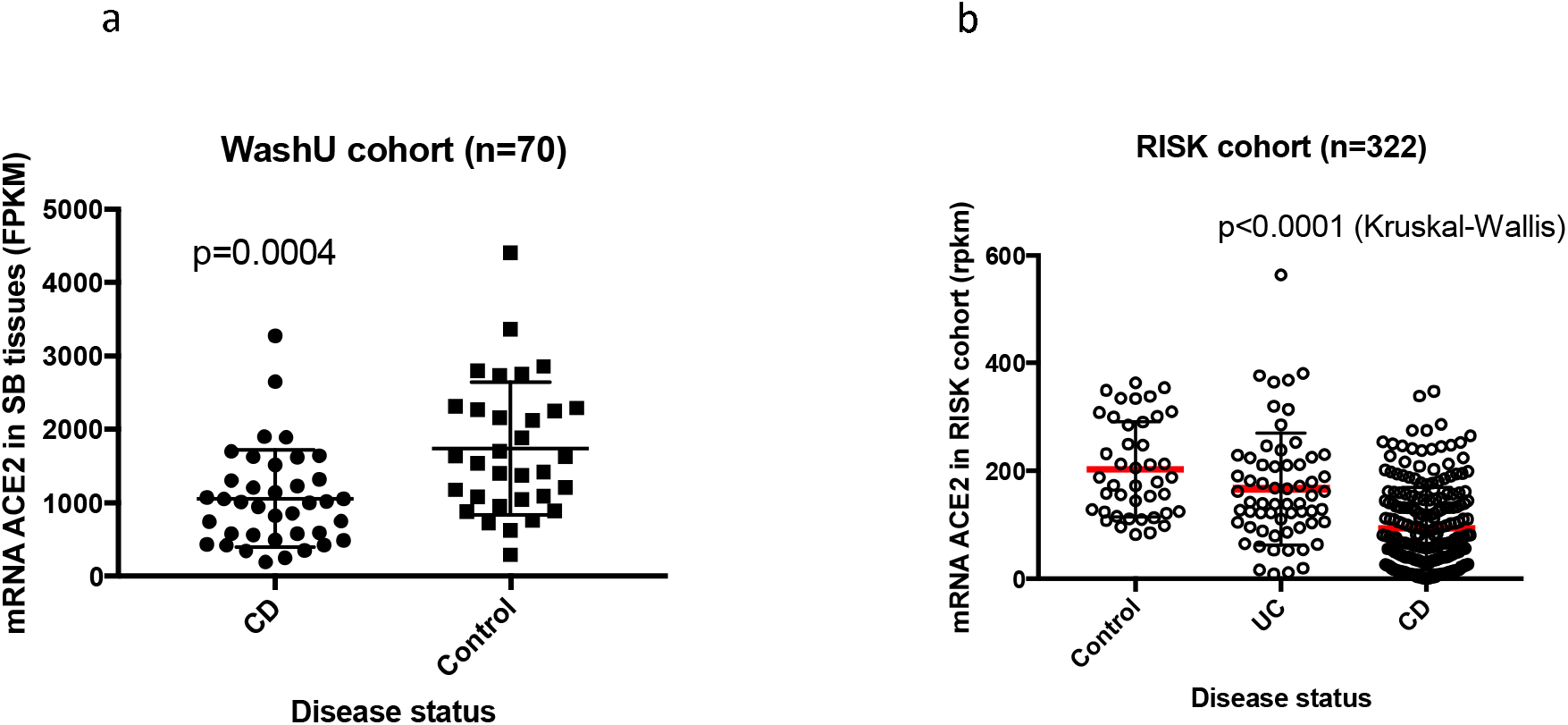
Association of *ACE2* with disease status; a) WashU cohort where ACE2 expression was downregulated in CD compared to controls (Mann-Whitney test, error bars indicate mean=+/-SD) ; b) RISK cohort where differences were seen in median *ACE2* expression in CD, UC and control (p<0.0001, Kruskal-Wallis, error bars in red indicate mean=/-SD).

In the RISK cohort, median *ACE2* expression in CD, UC, and control was statistically different (p<0.0001) (Figure 4b). Univariate models of *ACE2* expression with disease status indicated *ACE2* was lower in CD compared to controls (p=9.78e-14) as well as CD compared to UC (p=3.13e-09) (Table 3).

#### Multivariate associations

Multivariate models with disease status as predictor, were statistically significant or trending towards significance for lower *ACE2* expression in CD versus control in the WashU cohort (Table 2). In this cohort, we observed BMI as the strongest predictor of *ACE2* expression after adjusting for age at collection, disease status and gender. In the RISK cohort we observed lower *ACE2* expression in CD compared to controls (p=2.14e-14) or UC (p=5.3e-09) after adjusting for age at diagnosis and gender (Table 3). Age at diagnosis was significantly associated with *ACE2* expression after adjusting for disease status and gender in the RISK cohort (Table 3).

### Differences in small bowel *ACE2* gene expression by disease sub-phenotype

In the RISK cohort, where biopsies were taken from macroscopically uninflamed tissue, small bowel *ACE2* expression was lower in CD with small bowel involvement (iCD) in contrast to CD where the small bowel was un-involved CD (colonic CD) (cCD) (p=0.005) (Figure 5a, Table 4). Median *ACE2* expression was statistically different in controls, UC, iCD and cCD (p<0.0001) (Figure 5a). We also found a trend towards association of *ACE2* expression at diagnosis with the development of complicated disease by year 3 both without and with adjustment for age and gender (Figure 5b, p=0.076). This association of *ACE2* expression at diagnosis and subsequent development of complicated disease became significant by year 5 (Figure 5b, B2+B3 versus B1, p=0.017 and B2 versus B1, p=0.007; after adjusting for age and gender).

**Figure 5:**
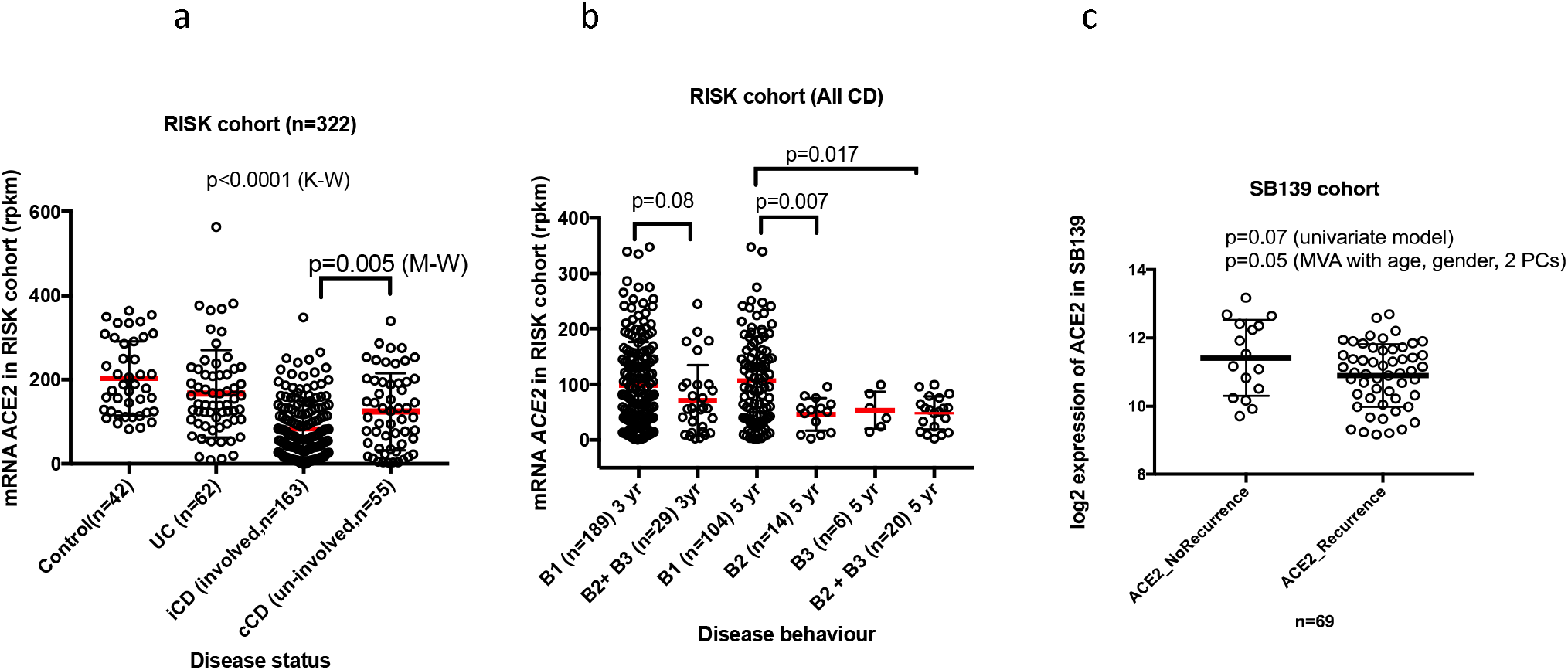
Association of *ACE2* with disease sub-groups; a) In the RISK cohort, *ACE2* expression was lower in involved CD (iCD) compared to un-involved CD (cCD) (p=0.005, Mann-Whitney test). Median *ACE2* expression was statistically different in Control, UC, iCD and cCD (p<0.0001, Kruskall-Wallis test, red bars indicate mean) ; b) In the RISK cohort, *ACE2* expression at diagnosis was lower in patients who developed complicated disease (B2 (stricturing) + B3 (penetrating) by year 3 compared to patients who remained uncomplicated (B1 (inflammatory)) at 3 year follow-up (red bars indicate mean). This association became significant by year 5 after diagnosis (B2+B3 versus B1, p=0.017; B2 versus B1, p=0.007, adjusted for age and gender); c) In the SB139 cohort, lower *ACE2* gene expression was observed in subjects with disease recurrence after surgery (p=0.05, adjusted for age, gender and 2 principal components (PCs)).

We previously reported that transcriptomics-based sub-groups with varying disease severity in the SB139 cohort where we found a severe refractory sub-group (CD3) was associated with increased recurrence, faster time to both recurrence and second surgery compared to the ‘mild’ refractory (CD1) sub-group [13]. We found that gene expression probe for *ACE2* was lower in the CD3 versus the CD1 sub-group (FC=-3.23, corrected p < 1e-07). In the SB139 cohort, we also found lower *ACE2* gene expression in subjects with disease recurrence after surgery after adjusting for age and gender and first two principal components in genotype data. (Figure 5c, p=0.05)

### *ACE2* in relation to other COVID-19 implicated genes, inflammatory cytokines, and known IBD targets

Due to the role of *ACE2* in COVID-19, we also examined the differential expression of COVID-19 related genes *ACE, TMPRSS2* and *SLC6A19* in controls versus CD in WashU (Table S2) and RISK cohorts (Table S3). Similar to *ACE2*, expression of *ACE* was lower in CD versus non-IBD controls in both WashU and RISK cohorts (Table S1 and S2). We found greater expression of the protease, *TMPRSS2* in CD compared to non-IBD controls while *SLC6A19* was lower in CD compared to controls in the RISK cohort (Table S3).

In the *ACE2* co-expression analysis we observed several genes that correlated with *ACE2* expression in both SB139 and the Cedars100 CD cohorts (Table S4) including *SIGMAR1* (coefficient = 0.348 to 1.55, p<0.0001) and *JAK1* (coefficient=1.51 to 1.74, p<0.05). *JAK3* was inversely correlated with *ACE2* (coefficient = -0.939 to -0.671, p<0.001) in both CD cohorts (Table S4).

### The effect of anti-cytokine therapy on *ACE2* expression

We performed univariate analyses for cohorts where small bowel samples were collected pre- and post-exposure to anti-TNF (infliximab) and anti-IL12/23 (ustekinumab) (IFX and UST cohorts) to query the effect of anti-cytokine monoclonal antibodies used in the treatment of IBD on small bowel *ACE2* expression. In the UST cohort we observed a trend towards increased *ACE2* expression between pre-treatment (week 0) and post-treatment (week 6) samples in the inflamed but not uninflamed tissues (Figure 6a). In the UST cohort meta-data for response to treatment was unavailable. In the IFX cohort, *ACE2* expression significantly increased after infliximab induction (p=0.02). This phenomenon was statistically significant in individuals who responded to treatment (p=0.037) but not in non-responders (Figure 6b).

**Figure 6:**
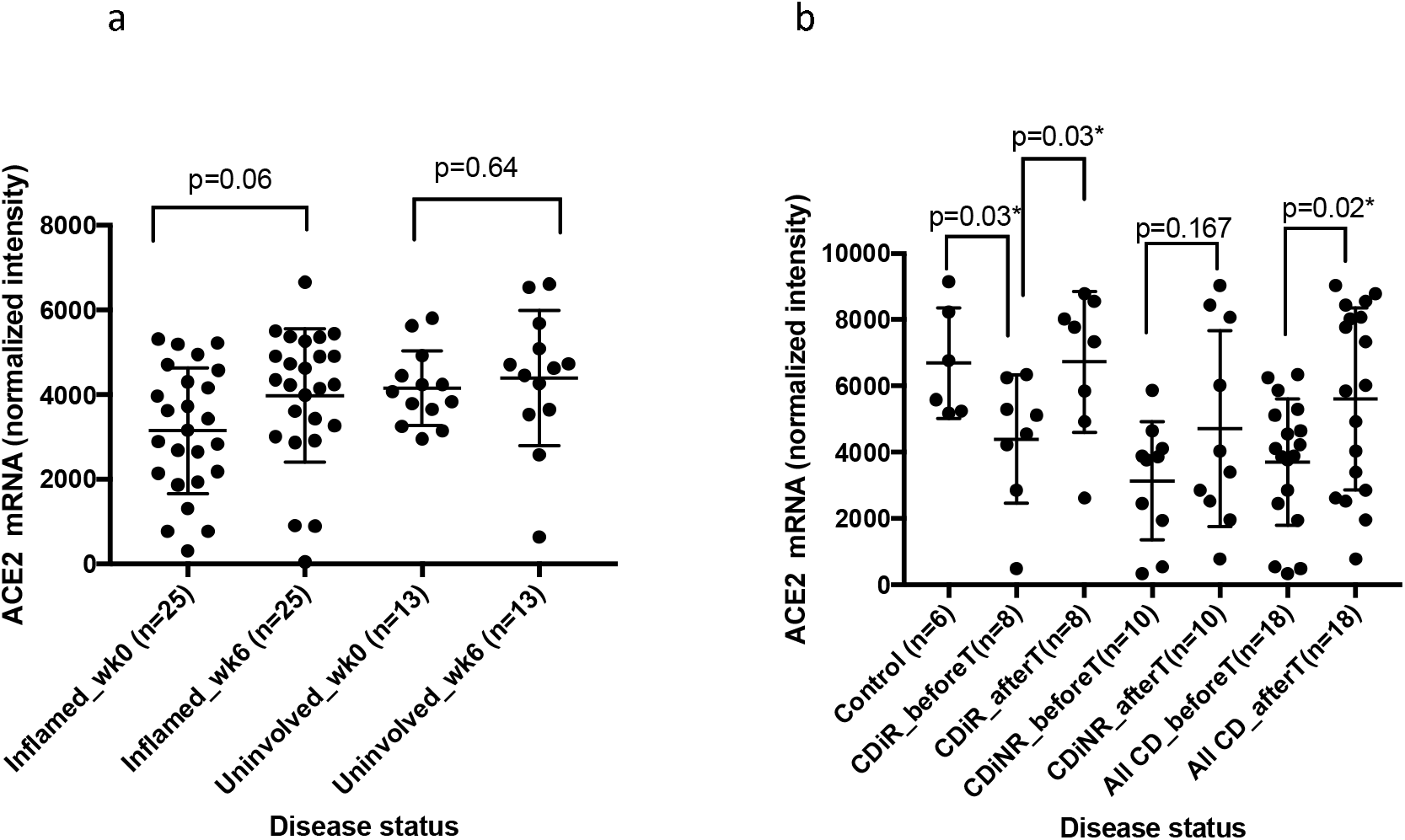
Univariate analysis, *ACE2* and IBD medication: a) *ACE2* levels in UST cohort in ileal inflamed samples before (week (wk) 0) and after (wk 6) (p=0.06, t test). b) *ACE2* levels in IFX cohort in controls (i.e. non-CD cases not treated with infliximab) are significantly higher (p=0.03, t test) than in Crohn’s ileitis responders before (CDiR_before) treatment. Six weeks after (CDiR_after) infliximab treatment the levels are significantly restored in responders compared to before treatment (CDiR_before) (p=0.03, t test). No significant difference was seen in Crohn’s ileitis non-responders before (CDiNR_before) and 6 weeks after (CDiNR_after) infliximab treatment.

## Discussion

Consistent with tissue and cell type specific expression of *ACE2* reported in the literature [22-24], we found robust expression of *ACE2* mRNA in small bowel ileal tissue from both non-IBD controls and subjects with CD. We observed evidence of increased *ACE2* mRNA in the ileum with demographic features that have been associated with poor outcomes in COVID-19 including age and elevated BMI. This age-related *ACE2* expression may be one of the reasons for decreased COVID-19 susceptibility in children versus adults if this data, particularly from the non-IBD subjects, are reflective of *ACE2* expression in other organs such as the lung [25]. Lower *ACE2* expression in uninflamed small bowel tissue is associated with recurrence after surgery and remarkably, from tissue taken at diagnosis, the subsequent development of complicated disease at 5 years after diagnosis in CD. These associations in non-IBD subjects and also the relationship between *ACE2* expression in macro- and microscopically non-inflamed tissue from CD patients point to systemic changes influencing *ACE2* mechanisms. In the cases of aging and increased BMI, both conditions are associated with increased immune tone and myeloid skewing [26, 27] as well as increased *ACE2*. Increased *ACE2* expression in lung has also been reported to be associated with age [28]. There is speculation that the GI tract may serve as an alternate route for uptake of SARS-COV-2 and our findings in the GI tract may take on increased relevance if this is confirmed [3, 23, 29]. Furthermore, early, but uncontrolled, evaluations of the SECURE-IBD registry [30] suggest that patients with IBD appear to be under-represented in those diagnosed with COVID-19 compared with what has been seen in the general populations in both Northern Italy and China [31]. Our data suggesting reduced *ACE2* expression in subsets of IBD may potentially contribute to this phenomenon, although further work needs to be done to better understand these observations.

Recent findings have suggested that men have a significantly higher mortality rate in COVID-19 but we did not observe higher *ACE2* expression in men – in fact in one cohort we observed higher expression in women. This finding is in keeping with *ACE2* expression in women from GTEx [12]. However, gender differences in *ACE2* expression may be tissue dependent and reflect tissue-specific escape from X-inactivation [32]. Differences in *ACE2* enzyme activity secondary to the effect of sex hormones has also been described [33]. Whether men are more susceptible to COVID-19, more likely to experience worse outcomes, or both, remains unknown. We saw a trend towards increased *ACE2* expression in smokers in only one of our cohorts, perhaps reflecting limited power given the relatively low frequency of smokers in our populations.

*ACE2*, which is part of the Renin-Angiotensin-Aldosterone System (RAAS), has been speculated to play a paradoxical role in disease progression of COVID-19 [22, 34, 35]. Although higher expression of *ACE2* increases viral uptake by host, physiologically *ACE2* has a significant anti-inflammatory role. This paradox is summarized in Figure 7 as it relates to CD and COVID-19. *ACE2* expression is required to neutralize the pathological effects of increased Angiotensin-II (Ang-II) in the classical RAAS pathway by converting Ang II to Ang1-7. Lung *ACE2* expression is protective against diseases such as pulmonary fibrosis, lung injury, and asthma [36]. Consistent with the reported literature of the anti-inflammatory role of *ACE2*, we report that within CD, reduced small bowel *ACE2* expression was associated with inflammation, more severe disease, and higher rates of disease relapse (even when measured in non-inflamed tissue).

**Figure 7:**
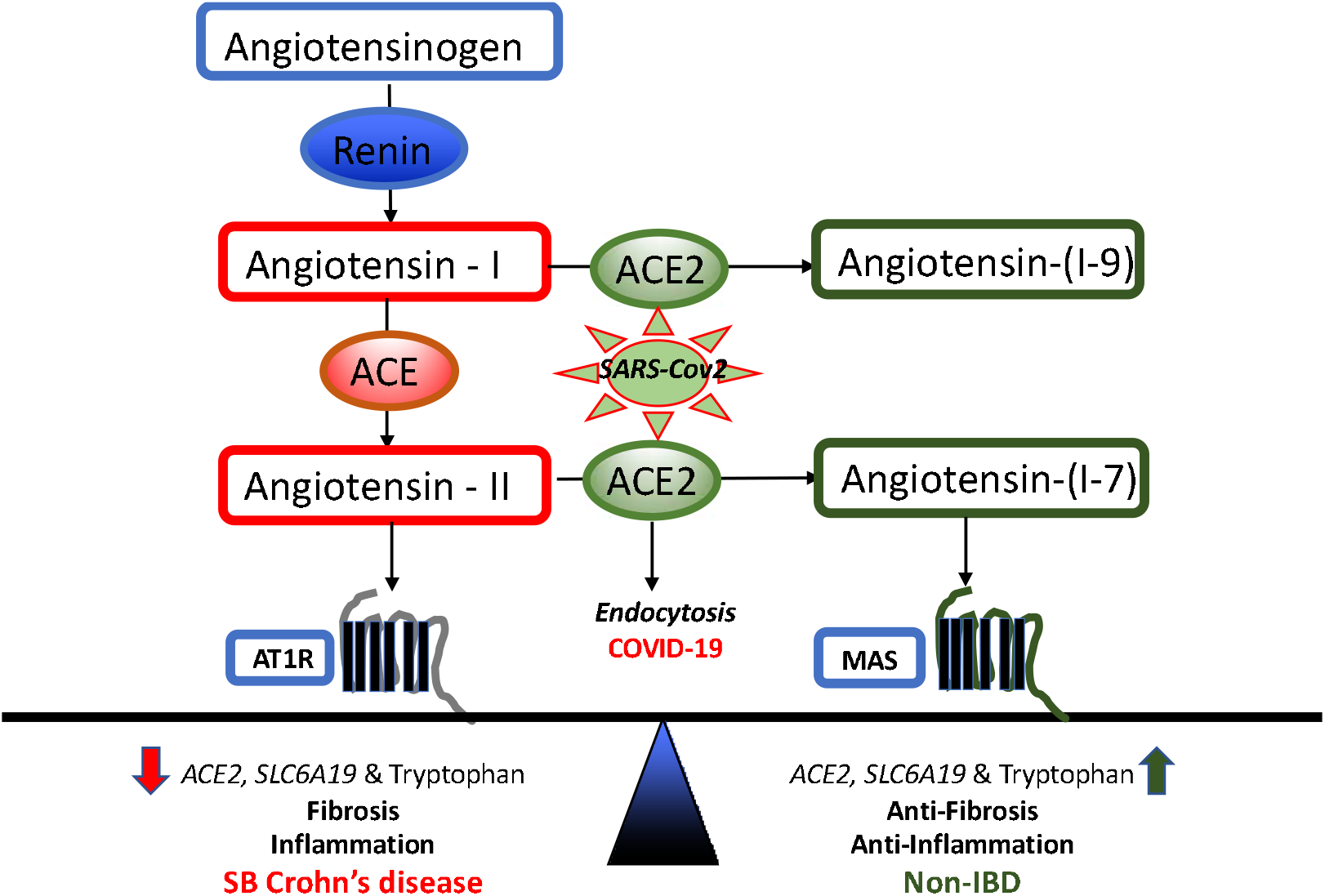
Main components of RAAS pathway active in small bowel (SB) Crohn’s disease (left) compared to non-IBD (right). We hypothesize that *ACE2* downregulation in CD leads to low expression of *SLC6A19* and hence, low tryptophan, tipping the balance towards an axis of inflammation and fibrosis. Restoration of *ACE2* levels leads to homeostasis which can be disrupted by SARS-COV-2 virus uptake in COVID-19. This figure is adapted from prior literature on RAAS pathway (11,35,41,42). AT1R: Angiotensin 1 receptor, MAS: MAS receptor.

How might downregulation of *ACE2* in severe CD lead to inflammation and intestinal injury? *ACE2* expression in the gut is necessary to maintain amino acid homeostasis, antimicrobial peptide expression and a ‘healthy’ intestinal microbiome and *Ace2*^*-/-*^ mice are more prone to developing colitis in induced models [9, 22, 37]. Expression of amino acid transporter *SLC6A19* (B(0)AT1) in SB is dependent on presence of *ACE2* [38] and *ACE2* acts as a chaperone for membrane trafficking of *SLC6A19* [39]. Accordingly, we find that expression of *SLC6A19* is decreased in CD along with that of *ACE2*. Notably, lower *SLC6A19* levels are selectively associated with lower tryptophan levels in small bowel CD [37, 40] (Figure 7, adapted from prior literature on RAAS pathway [11, 35, 41, 42]). Dysregulated tryptophan metabolism has been linked to systemic inflammation [43, 44]. The biologic mechanisms that link levels of tryptophan to disease intestinal inflammation and obesity are complex, including host and microbial production of bioactive tryptophan metabolites [40, 45, 46] and the selective roles of these metabolites on molecular processes such as energy checkpoint [37, 47, 48] and transcriptional controls of inflammation pathways [49, 50]. Exploring these mechanisms in the *ACE2* deficiency of CD may distill how the *ACE2* network could serve as a protective pathway for IBD. Intriguingly, COVID-19 complications may be more common in IBD patients with active disease and studying the role that *ACE2* mechanisms play in both acute infection and complication must be a priority for the research community.

Elevated *ACE2* levels may promote tissue propagation of virus and, in theory, could promote COVID-19 disease severity. However, the secondary cytokine storm likely promotes tissue injury via mechanisms independent of viral propagation and this process may be independent of *ACE2* expression. Alternatively, *ACE2*, with its anti-inflammatory properties may play a role in protection from the secondary cytokine storm. Due to the SARS-CoV-2/*ACE2* interaction, there has been interest in treatments for COVID-19 that modulate *ACE2*. A study examining *ACE2* with TNF-α production in SARS-CoV found that viral entry modulated TNF-α-converting enzyme (TACE) via the *ACE2* cytoplasmic domain and caused tissue damage through increased TNF-α production [51]. We identified that *ACE2* levels were restored after infliximab therapy and that this was statistically significant in anti-TNF responders. We also observed an increase in *ACE2* expression after ustekinumab therapy but only in inflamed tissue although this change did not achieve statistical significance. The inverse relationship of *ACE2* with inflammatory cytokines and restoration of enhanced *ACE2* levels after response to anti-TNFs point towards the anti-inflammatory function of *ACE2*. However, further work will be needed to delineate this relationship and determine whether anti-TNFs could be effective in modulating the secondary cytokine storm associated with COVID-19 [52].

Finally, we examined *ACE2* co-expression with a set of candidate genes as potential targets for novel or repurposed drugs. For example, our analysis revealed *SIGMAR1* to be consistently co-expressed with *ACE2* in two cohorts. *SIGMAR1* has been identified as a candidate target for the drug hydroxychloroquine and several clinical trials are testing the efficacy of hydroxychloroquine to treat COVID-19. In addition, we identified that *JAK1* expression is consistently correlated with *ACE2* expression in contrast to *JAK3* which shows a consistent but inverse relationship with *ACE2*. Selective JAK inhibitors are available and in development and baricitinib (a JAK1/2 inhibitor) is being tested in COVID-19 based on both its anti-inflammatory properties and its possible role in inhibiting endocytosis and viral entry [53]. Our observation of co-occurrence of *ACE2* and *JAK1* provides some support for the testing of this compound in COVID-19.

To summarize, we observed association of *ACE2* with various demographic and clinical factors in multiple ileal transcriptomic datasets. Our associations are largely consistent with demographic criteria associated with worse outcomes from COVID-19. We also report an impaired *ACE2* gene expression that leads to worse outcomes in CD and evidence that implicates *ACE2* pathway as a protective, tryptophan-dependent anti-inflammatory mechanism in severe IBD. Anti-TNF and perhaps anti-IL12/23 may restore *ACE2* levels in the context of inflammation reduction, suggesting that restoration of the *ACE2* pathway may be a mechanism by which these drugs promote recovery in IBD. Our work supports the potential paradoxical function of *ACE2* in inflammation and COVID-19 infection. Individuals with higher *ACE2* expression may be at increased infection with SARS-CoV-2 but *ACE2* likely has anti-inflammatory and anti-fibrotic functions and may play an important role in preventing the secondary cytokine storm seen in some individuals with COVID-19 as well as preventing the development of complicated disease in IBD. How TNF-α and anti-TNF therapies as well as other immune targeting therapies may modulate these processes requires urgent investigation.

## Data Availability

Transcriptomics datasets used in this manuscript are available on public dataset (GEO, Accession #s: GSE120782, GSE57945, GSE16879,GSE100833). One is available at arrayExpress (E-MTAB-5783). One of the dataset is unpublished and will be deposited on GEO.

## Acknowledgements

We are thankful to all clinicians, coordinators and especially patients who have contributed time, data, and samples to the MIRIAD Biobank.

